# Trend Analysis on the Application of Social Work Professionalism: The Case of Refugee Child-Focused Humanitarian Organizations in Ethiopia

**DOI:** 10.1101/2024.10.28.24316172

**Authors:** Fisehatsion Afework, Mesele Mengsteab

## Abstract

This study investigated the trend in the application of social work professionalism in delivering child-focused social work services for refugee children within Ethiopian refugee camps. The study seeks to understand the how the various constituents of social work professionalism has been applied by child-focused humanitarian organizations between the years of 2017 to 2023. A time series trend analysis model employed along with a mixed-methods approach across three refugee camps selected through multi-stage cluster sampling. The research investigates how social work services align with professional standards and the challenges faced by practitioners. Findings reveal significant caseloads, resource constraints, and ethical dilemmas in service prioritization. However, positive trends emerge, including the implementation of the Child Protection Minimum Standards framework, a client-centered approach, and growing collaboration with the families of refugee children. The study translates these findings into actionable recommendations for strengthening social work practice, including increased funding, caseload management strategies, prioritizing child participation, and fostering inter-agency collaboration. A proposed model generated through the study visually depicts key challenges, positive trends, and recommendations to improve child-focused social work services through the lens of professional ethics in refugee settings. This research underscores the importance of a multi-pronged approach to ensure high-quality, ethical services for all refugee children.

## Introduction

Child-focused humanitarian organizations are specialized agencies providing child protection and other social work services tailored to vulnerable children who are in a protracted crisis context. Initially, child-focused social services were delivered through charitable organizations with less emphasis on professional standards. Following World War II, the social work profession gained stability and established core values like service, social justice, and dignity of the person. This led to a more professionalized approach to social work within humanitarian organizations, with social workers adhering to ethical codes and specialized practices, particularly when dealing with vulnerable populations like children in crisis (Alliance for Child Protection in Humanitarian Action, 2023).

Ethiopia faces challenges in defining the role of social workers within healthcare and other settings, particularly regarding HIV/AIDS interventions (Hailu, 2013). Furthermore, social workers employed by various civil society organizations (CSOs) often lack the practical skills and experience required for agency-level work (I-TECH, 2010). This trend extends to child-focused humanitarian organizations in Ethiopia. These organizations often rely heavily on unqualified refugee incentive workers to deliver child-focused social work services. As an example, in 2014, a total of 152 para-social workers (refugee incentive workers) were responsible for providing entire child protection case management services in Gambella refugee camps (UNHCR, 2014).

Despite legal instruments with indirect implications for social work practice in Ethiopia, there are no specific regulatory frameworks governing the development and management of the profession (Hailu, 2013). This lack of regulation allows service providers and non-governmental organizations to hire individuals who work directly with clients without any relevant professional qualification. Although social work professionalism is well-studied within traditional Human Service Organizations (HSOs), a significant gap exists in understanding its application within child-focused humanitarian organizations that serve refugee children.

This study proposed a unique approach by employing a trend analysis to investigate how social work professionalism has evolved within Ethiopian humanitarian organizations over a seven-year period (2017-2023). By focusing on Ethiopia, a country with a large refugee population, this research addresses a critical knowledge gap concerning social work professionalism in this specific context. This approach allows us to examine the changing nature of social work professionalism within these child-focused humanitarian organizations across time while delivering child-focused social work services and offering valuable insights to improve social work practices in refugee settings.

Currently, a gap exists in our understanding of how these organizations approach social work interventions with refugee children. This study aims to analyze a time-series trends towards the application of various constituents of social work professionalism across the years 2017 to 2023.

The year 2017 serves as the starting point due to the significant increase in refugee numbers associated with the renewed conflict in South Sudan (July 2016). This period also coincides with the opening of Nguenyyiel refugee camp in Gambella (October 2016), which currently hosts the largest refugee population in Ethiopia (UNHCR, 2020). Finally, the selection of 2023 as the endpoint ensures access to complete data, as refugee data in Ethiopia is compiled annually.

Through this trend analysis, the research aims to enhance our understanding of social work professionalism in child-focused humanitarian organizations operating in Ethiopia and serving refugee children.

### Justification for Time Period

The selection of the 2017-2023 as a timeframe for studying trends in the application of social work professionalism is justified by three key parameters

1. **Increased Refugee Influx:** 2017 witnessed the highest refugee influx in Ethiopia due to the South Sudanese conflict in July 2016.
2. **Opening of Major Refugee Camp:** This timeframe coincides with the opening of the largest refugee camp in Ethiopia, Nguenyyiel, in October 2016.
3. **Data Availability:** Refugee data in Ethiopia is compiled annually; selecting 2023 as the endpoint ensures access to complete data for the chosen timeframe.

### Constituents of social work professionalism

The study considered four major constituents of social work professionalism to frame the entire subject of analysis (i) types of child-focused services being provided by humanitarian organizations considering social work practices dimensions i.e., therapeutic, transformational, and social order dimensions (Payne, 2006), (ii) application of social work values and ethical principles i.e., NASW’s core values and codes of ethics, (iii) how social work practice interacts with expectations and/or regulations of those child-focused humanitarian organizations, and (iv) the qualification and professional development of social workers within those child-focused humanitarian organizations (Payne, 2006). The purpose of my research is to analyze a seven-year trend in the application of those constituents of social work professionalism between the year 2017 and 2023. The below key questions were rigorously answered at the end of the study

The below key questions were rigorously answered at the end of the study:

**1.** What types of child-focused social work services have been provided by humanitarian organizations for refugee children in Ethiopia since 2017?
**2.** How do the delivery format and quality of social work services offered by child-focused humanitarian organizations for refugee children in Ethiopia since 2017 changed across time to 2023.

## Methods

Throughout the data collection, analysis, and interpretation process of this study predominately a *‘positivist’* approach was applied. This is associated with a purely quantitative research design employed involving time series analysis of trend in several variables of social work professionalism and a subsequent trend forecasting for similar variables based on past trend records assuming any external factors affecting the prediction are constant. However, a lesser extent of *‘interpretivist’* approaches was used in a few component of this study. For instance, (i) a systematic technique used to synthesize the findings acquired from the quantitative analysis against few selected social work theories, (ii) a minimal level of subjective thoughts of respondents used with the purpose of complementing very few quantitative findings to tailor with the context at the ground level, and (iii) a document analysis conducted to learn how the NASW core values which is the guiding framework of social work practice in the academia is aligning with the child protection minimum standard in humanitarian action (CPMS) applicable at grassroots level.

To see how things have changed over time, a time series analysis of data employed for the data collected from 2017 to 2023. This helped to identify trends in the way social work services were provided. Surveys, key informant interviews, focus group discussion and document reviews were the major data collection mechanisms applied.

The study focused on 14 child-focused humanitarian organizations that work on refugee children in Ethiopia. The study looked at social workers, child protection officers, program managers, and other staff who are involved in providing child-focused social work services. Since the organizations were spread out across different refugee camps, the researcher used a multi-stage cluster sampling technique. This means they randomly choose a few regions and camps to focus on, and then randomly selected organizations and social workers within those areas.

Out of the 14 child-focused humanitarian organizations three child-focused humanitarian organizations were considered in this study who were selected based on a firm sampling procedures applied, (i) Innovative Humanitarian Solution (IHS), which is operating in Asayita refugee camp located in Afar regional state. IHS involved in providing child protection in emergency and multi-purpose cash assistance services for Eritrean refugee children hosted in Asayita refugee camp, (ii) Plan International Ethiopia (PIE) which is operational in Ngunyyiel refugee camp Gambella regional state (the biggest refugee hosting camp in the country) providing child protection and education in emergency services for South Sudanese refugee, (iii) at the third level Save the Children International (SCI) is also participant of this study which is also involved in child protection in emergency and early childhood and development services in Terikidi refugee camp within Gambella regional state.

This study used a multi-stage cluster sampling technique for the quantitative design because it was impractical to reach every social worker in all the refugee camps. Here’s how it worked:

- First, randomly picked two regions (clusters) out of five.
- Then, randomly selected refugee camps within those regions.
- Since only one organization operates per camp, all social workers from those organizations were included (census approach)

While a total of 150 social workers participated as respondents in the survey, 48 (8 Female) respondents who were selected purposively participated in focus group discussion and 6 interviewees took part in the key informant interviews.

### Time series analysis

was used to identify trends in the provision of child-focused social work services over the seven years (2017-2023). Depending of the identified nature of trend an appropriate time series models has been used to analyze current trends and predict upcoming trends until 2028. In the context of this study an ARIMA (Autoregressive Integrated Moving Average) and exponential smoothing models were the predominant time series models used.

### Regression analysis

was used to explore relationships between different variables associated with the constituent of social work professionalism for instance a chi-square test was applied to identify how those variables are associated with the increasing time period between 2017-2023.

## Findings

The tabular presentation below illustrates the demographic age and gender distribution of respondent social workers across three organizations i.e., Innovative Humanitarian Solution (IHS), Plan International (PI), and Save the Children International (SCI) The gender distribution demonstrates that HIS has a higher percentage of females (13.5%) compared to males (3.1%), while in PI females make up the vast majority of employees (82.7%) compared to males (81.6%), and SCI has similar trend to HIS which has a higher percentage of females (3.8%) compared to males (15.3%). Across all three organizations the average employee age is around 25.6 years old. The median age is 28 years old, indicating that half of the employees are younger than 28 and the other half are older.

**Table 1.**
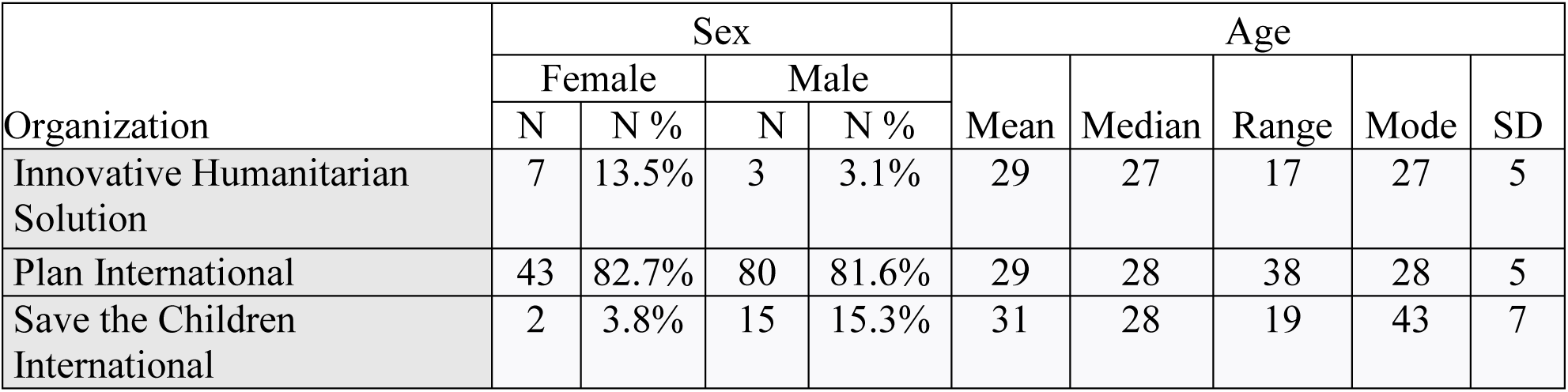
Demographic profile of respondents.

Over half (51.3%) of sample social workers only has a high school certificate as their highest level of education, and a quarter (24.7%) of the sample has a bachelor’s degree. A smaller fraction (19.3%) of has diploma, and a very insignificant proportion (1.3%) has a master’s degree. A very small percentage has only completed primary school or is illiterate.

**Figure 1.**
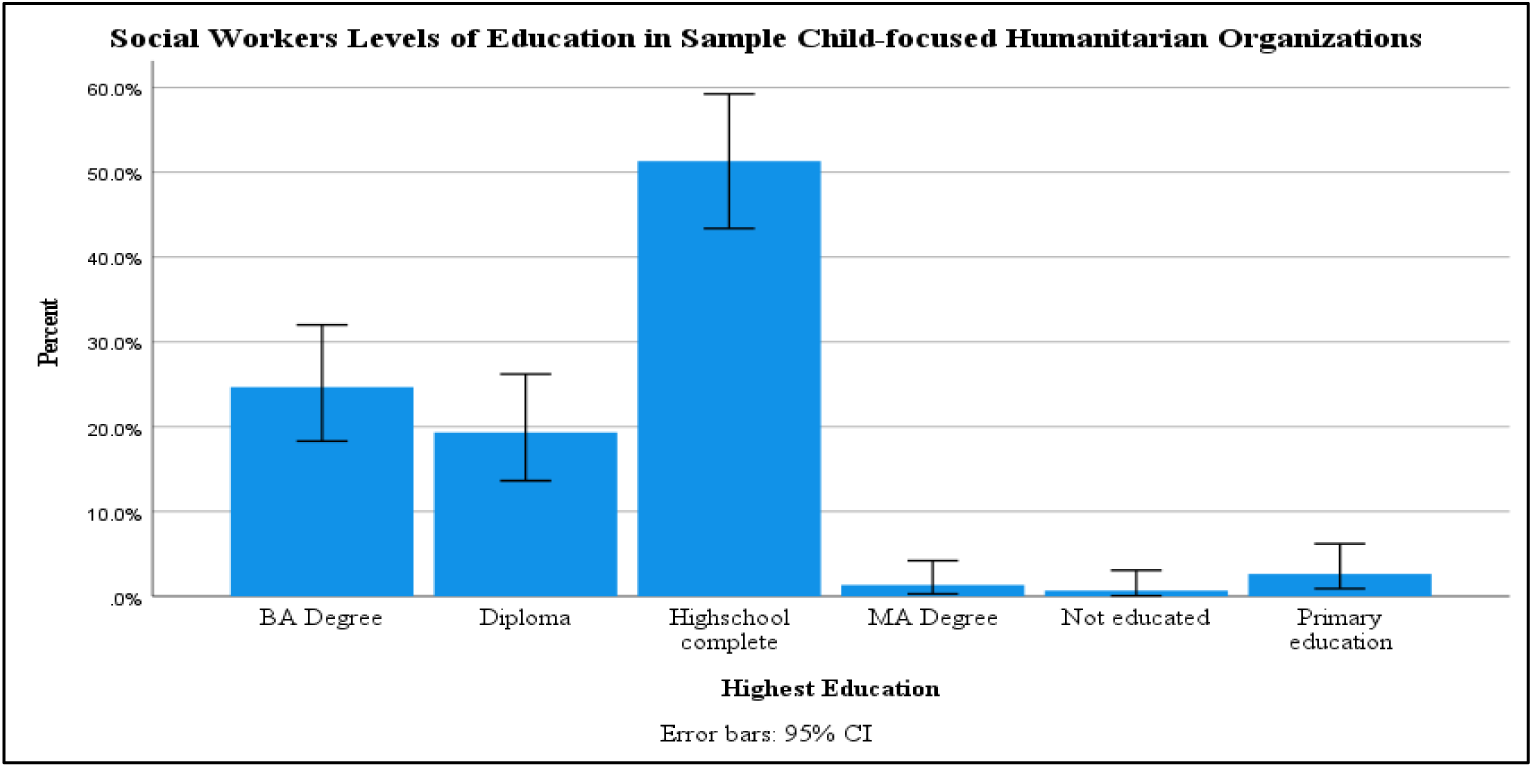
Social Worker Levels of Education.

On the other hand, 54 % of the social workers providing child-focused social work services for refugee children are primary and/or high school education completed (54%). Only 14 % of them have studied social work by academic training and 14% also accounts for nursing field of study, followed by other fields like sociology and psychology (around 5% each). Public health and medicine have a smaller presence (2% and 0.7% respectively). There’s one social worker listed as illiterate. Furthermore, for the interest of this research a cross-tabulation analysis was executed in order to investigate in which areas of child-focused social work services those social workers by academic training are involved. The cross-tabulation result indicates that only 15 % of the delivery of the above 6 (six) areas of child-focused social work services covered only by 15% by professional social workers by academic training. Particularly social workers by academic training are highly involved in overall child protection service and psychosocial support. The rest majority portion of the services is covered by high school graduates and primary school complete incentive workers (56%) and with little involvement of workers with nursing (14%), Sociology (4%), and Psychology (3%), Education (4%), and very insignificant proportion (2%) public health field of study background.

### Areas of child-focused social work services for refugee children

Based on a linear frequency analysis conducted while all of the respondent social workers are involved in providing overall child protection services, 31.3% have mentioned also that psychosocial support service is also among the child-focused social work services being provided for refugee children which is followed by family reunification (17.3%) and education services (14%).

**Table 2.** Areas of Child-focused services being provided for refugee children.

| Types of Service Frequencies |  |  |  |  |
| --- | --- | --- | --- | --- |
|  |  | Responses |  | Percent of Cases |
|  |  | N | Percent |  |
| Area(s) child-focused of social work services <sup>a</sup> | Education | 21 | 8.3% | 14.0% |
|  | Health including mental health | 1 | 0.4% | 0.7% |
|  | Family reunification | 26 | 10.3% | 17.3% |
|  | Overall child protection | 150 | 59.3% | 100.0% |
|  | Psychosocial support | 47 | 18.6% | 31.3% |
|  | Livelihood | 5 | 2.0% | 3.3% |
|  | Other | 3 | 1.2% | 2.0% |
| Total |  | 253 | 100.0% | 168.7% |
| a. Dichotomy group tabulated at value 1. |  |  |  |  |

### Trend in the types of child-focused social work services provided

A survey questionnaire was administered against the social workers employed in the three organizations operating in three subsequent refugee camps. The survey collected a time-series data on eleven types of child-focused social work services provided since 2017 up to 2023.

**Figure 1.**
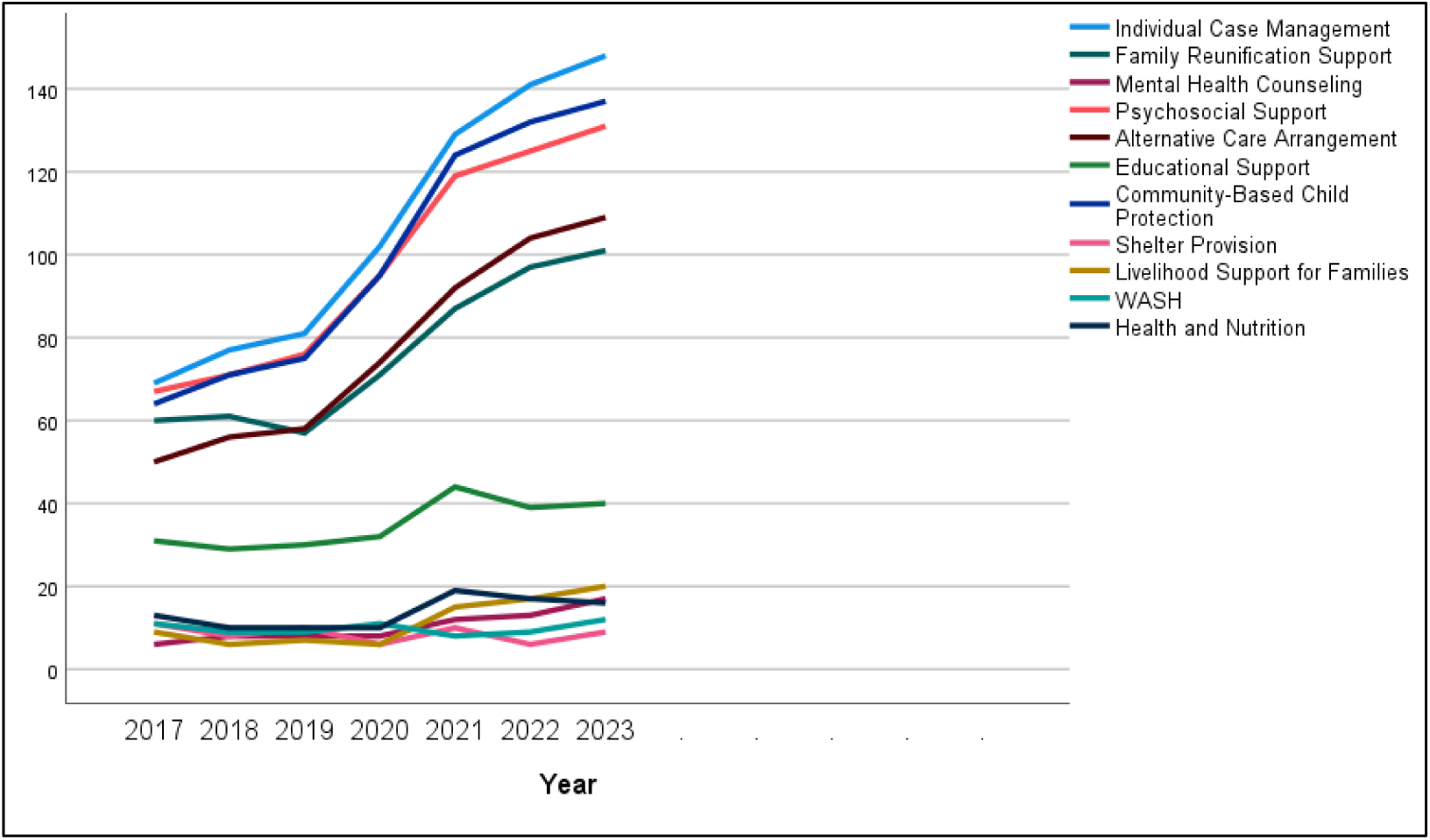
Trend chart for child-focused social work services for refugee children.

At initial level through plotting time-series sequencing chart the researcher was able to identify the nature of various trends in the provision of each child-focused social work services. Based on the preliminary sequencing chart three major trend characteristics of the services observed across the time series period.

***Trend-1:*** it is found that five (5) child-focused social work services (i.e., individual case management, community based child-protection, psychosocial support, alternative care arrangement, and family reunification) has a clear non-stationary and significantly increasing trend with consistently increasing means (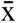).

***Trend-2:*** three (3) child-focused services (i.e., shelter provision, WASH service provision, and Health and nutrition) have demonstrated a random walk trend with no clear direction and no consistent decrease or increase of mean (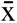).

***Trend-3:*** another three (3) child-focused services (i.e., mental health services, educational support, and livelihood support for families) showed insignificant increasing trend overtime and a little bit closer to stationarity of mean(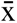).

A sequencing regression was applied to determine the nature of trend in those child-focused social work services when compared to ascending time period. R-square (R^2^) was calculated for each year against the frequency of each child-focused service types so as to measure the association between the trend and time. Based on the result the increasing trend of ‘Alternative care arrangement’ has the strongest association with the increasing time period (R^2^ = 0.921) which is followed by ‘Individual case management’ with (R^−^ = 0.914). However, the trend in providing child-focused service related to ‘Educational support’ and ‘Livelihood support for families’ has comparatively weaker association with the increasing time (R^2^ = 0.553 and R^2^ = 0.696). On contrary, the trend of two child-focused social work services for refugee children has insignificant association with the increasing time period (i.e., Shelter Provision and Water, Sanitation and Hygiene/ WASH).

A statistical test on the stationary nature of the trend of above eleven types of child-focused social work service was applied to decide the specific time series model for further analysis. According to stationary test Auto Regressive Integrated Moving Average (ARIMA) model was used to analyse the trend of the eight child-focused service type and Exponential Smoothing was utilized to analyse the trend for the remaining three child-focused social work services. As per the model fit statistical test result, the stationary R-squared value (R²ₐ against each dependent variables of child focused social work services (i.e., individual case management, alternative care arrangement, psychosocial support, community-based child protection, family and reunification support, shelter provision, WASH, and health & nutrition) indicates (R²ₐ= = 0.000) which qualities the ARIMA model as a best fit for the analysis of the trend. However, the remaining three child focused social work services has an approximate R²ₐ= 1. (i.e., mental health counselling, education support and livelihood support for families) which allowed the researcher to use exponential smoothing to analyse the trend across the study period (Shin, 2017).

**Table 3.** Parameters for time-series model service types.

| Exponential Smoothing (Holt) Model Parameters |  |  |  |  |  |  |
| --- | --- | --- | --- | --- | --- | --- |
| Model |  |  | Estimate | SE | t | Sig. |
| Mental Health Counseling | No Transformation | Alpha (Level) | 0.00 | 0.47 | 0.00 | 1.00 |
|  |  | Gamma (Trend) | 0.00 | 273.06 | 0.00 | 1.00 |
| Educational Support | No Transformation | Alpha (Level) | 0.20 | 0.26 | 0.75 | 0.49 |
|  |  | Gamma (Trend) | 0.00 | 0.31 | 0.00 | 1.00 |
| Livelihood Support for Families | No Transformation | Alpha (Level) | 0.00 | 0.63 | 0.00 | 1.00 |
|  |  | Gamma (Trend) | 0.00 | 364.80 | 0.00 | 1.00 |

| ARIMA Model Parameters |  |  |  |  |  |  |
| --- | --- | --- | --- | --- | --- | --- |
|  |  |  | Estimate | SE | t | Sig. |
| Individual Case Management | No Transformation | Constant | 13.167 | 3.664 | 3.593 | .016 |
|  |  | Difference | 1 |  |  |  |
| Family Reunification Support | No Transformation | Constant | 6.833 | 3.188 | 2.144 | .085 |
|  |  | Difference | 1 |  |  |  |
| Psychosocial Support | No Transformation | Constant | 10.667 | 3.499 | 3.048 | .028 |
|  |  | Difference | 1 |  |  |  |
| Alternative Care Arrangement | No Transformation | Constant | 9.833 | 2.638 | 3.727 | .014 |
|  |  | Difference | 1 |  |  |  |
| Community-Based Child Protection | No Transformation | Constant | 12.167 | 4.110 | 2.960 | .032 |
|  |  | Difference | 1 |  |  |  |
| Shelter Provision | No Transformation | Constant | 8.571 | .751 | 11.407 | .000 * |
| WASH | No Transformation | Constant | 9.857 | .553 | 17.816 | .000 * |
| Health and Nutrition | No Transformation | Constant | 13.571 | 1.429 | 9.500 | .000 * |

Based on the selected time-series model statistics a prediction in the future trends of each service types made between the years 2024-2028 taking in to account the historically observed trend between the years 2017 and 2023. The time series predication result indicates that three (3) among the eight child-focused services type (i.e., health and nutrition, WASH and Shelter provision services for refugee children) will have a statistically significant (sig. = .000) which will have stationary and unpredictable trend. However, the remaining eight (8) child-focused services will probably have an increasing and non-stationary trend based on similar experience in previous period.

Moreover, as a complement to the survey result, the FGD highlighted various services offered to support refugee children in the refugee camp. These include psychosocial support to help them cope with emotional difficulties. Basic necessities like clothing and hygiene products are provided through Non-Food Items (NFIs) for vulnerable children. Adolescents benefitted from life skills training to make positive choices in life, though this program has seen a reduction since 2022. Previously, adolescents also had access to a library and a Child-Friendly Space for recreation and learning, but these services have transitioned to support youth programs. Importantly, the program facilitates referrals for children requiring specialized care and arranges alternative care for abandoned children. Finally, they offer family tracing and reunification services to help children reconnect with their families if separated during displacement.

### Formats of delivery for child-focused social work services

A comparison of mean was conducted against the frequency of responses respective to each year and individual physical set up where child-focused services are provided. The result indicates that child friendly spaces are consistently considered as the most (35 %) preferred place by the sample humanitarian organizations to provide child-focused social work services. Schools and food distribution centers followed with 14 % and 13 % preference to provide child-focused social work services for refugee children.

**Table 3.**
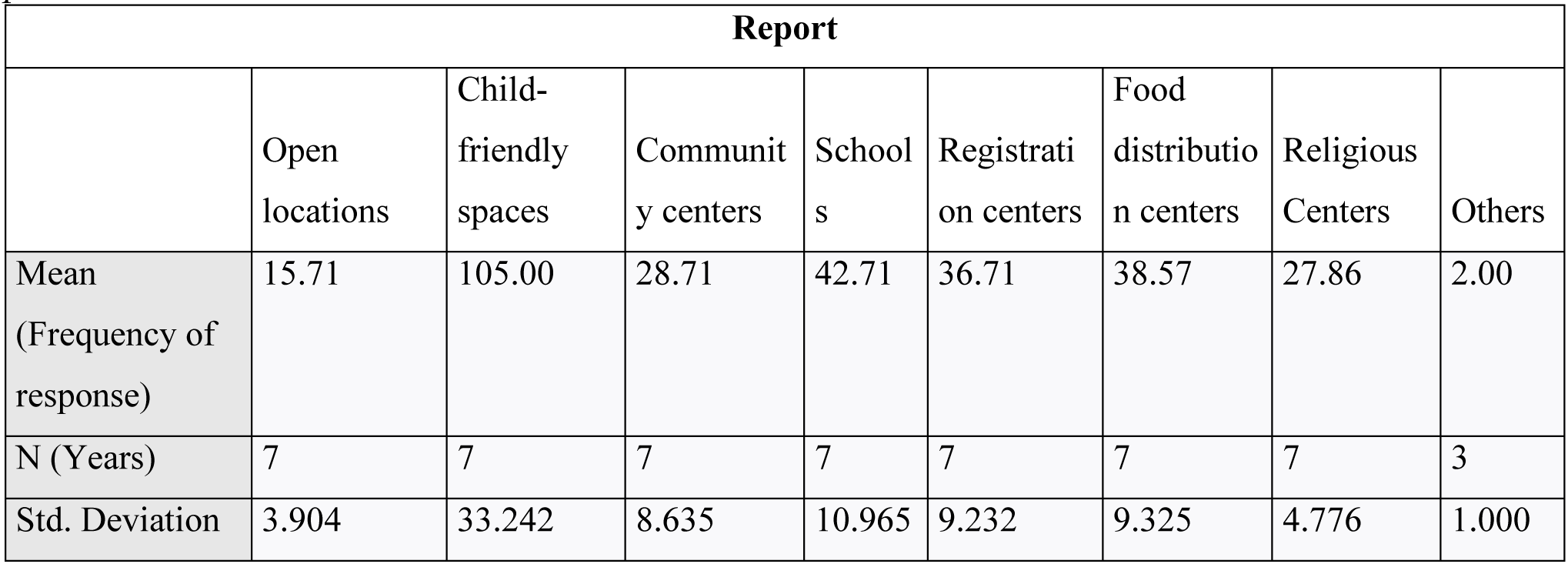
Mean Comparison on the places (settings) where child-focused social work services provided.

| Report |  |  |  |  |  |  |  |  |
| --- | --- | --- | --- | --- | --- | --- | --- | --- |
|  | Open locations | Child-friendly spaces | Community centers | Schools | Registration centers | Food distribution centers | Religious Centers | Others |
| Mean (Frequency of response) | 15.71 | 105.00 | 28.71 | 42.71 | 36.71 | 38.57 | 27.86 | 2.00 |
| N (Years) | 7 | 7 | 7 | 7 | 7 | 7 | 7 | 3 |
| Std. Deviation | 3.904 | 33.242 | 8.635 | 10.965 | 9.232 | 9.325 | 4.776 | 1.000 |

In addition a linear sequencing analysis was conducted to see trend in the place preference of humanitarian organizations to provide child-focused social work services. The sequencing analysis indicates child friendly space shows a non- stationary and significant increasing trend (R^2^ = 0.938) compared to other preferred places (settings) across the year from 2017-2023.

**Figure 2.**
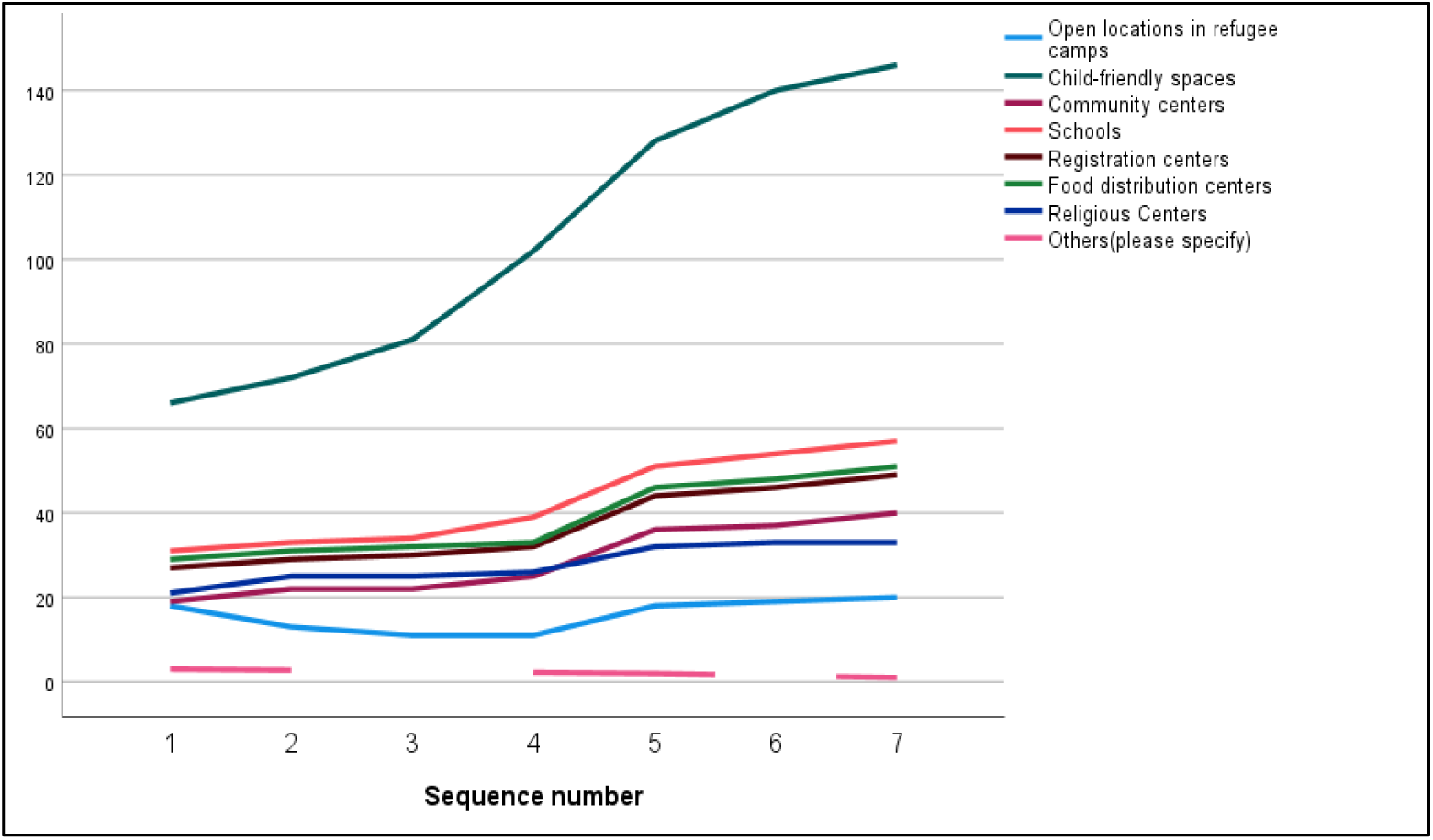
Trend of the place where child-focused social work services provided.

### Levels of child-focused social work interventions

Respondents were also asked about the various levels of intervention they have been using across time while providing child-focused social work services for refugee children between the years 2017-2023. The result demonstrated that; (i) individual level of intervention has a statistically significant increasing and non-stationary trend across time. Most importantly, its trend have strong association with years (R^2^ = .945), (ii) community level intervention also indicates that it have a consistently increasing trend with strong association with time or years (R^2^ = .922), (iii) family level intervention have also an increasing trend across time with a non-stationary trend and still strong trend association with time (R^2^ = .894), (iv) group work (level of intervention) has a very slight increasing trend and moderate association with time (R^2^ = 783), and lastly (v) intervention at policy or advocacy level has an insignificant increasing trend and stationary nature particularly starting from the year 2021. The R-squared result for policy level intervention showed a weak association with time (R^2^ = .526).

**Figure 3.**
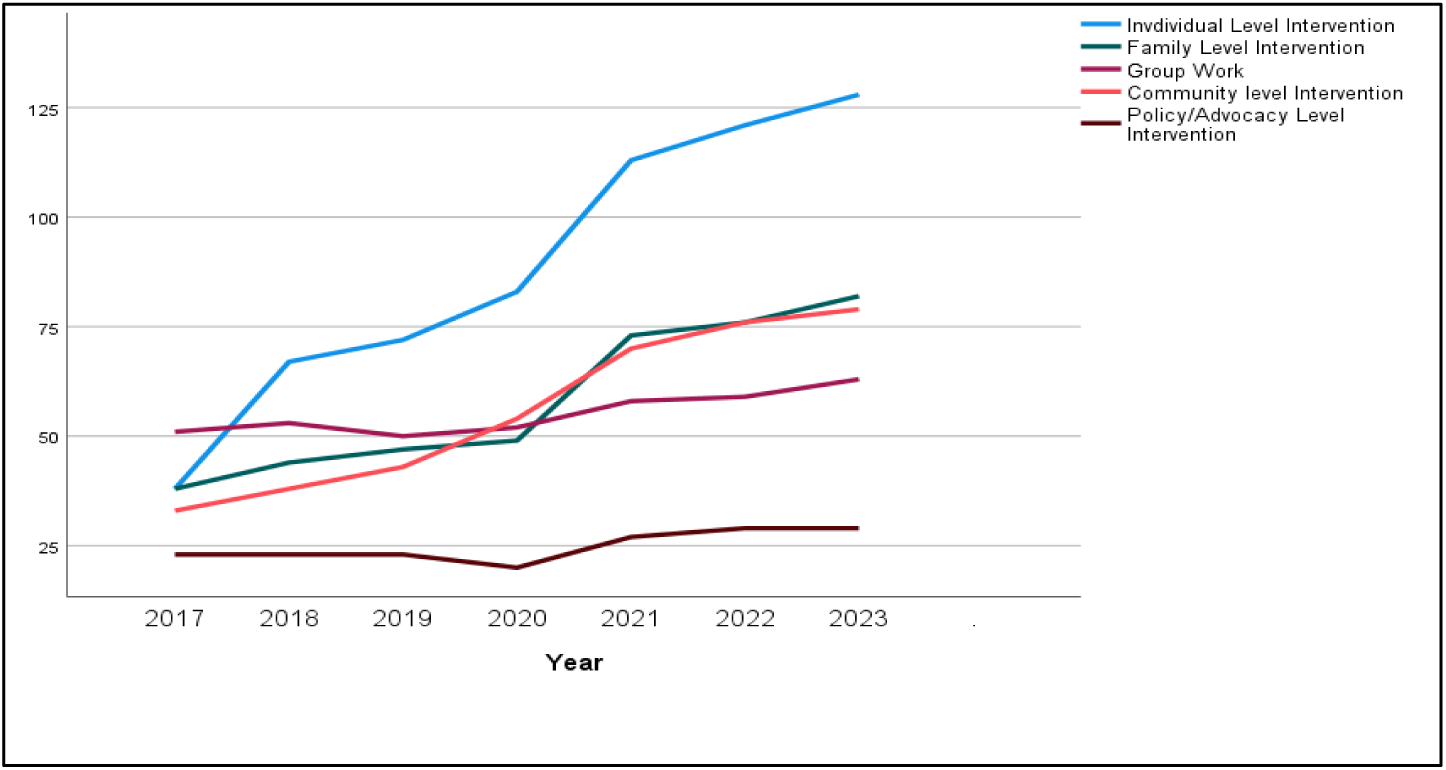
Trend on the level of interventions while providing social work services.

Based on sationarized R-square value (R^2^_a_), three levels of interventions (i.e., individual, family, group and policy level has demonstrated an R^2^_a_ value closer to 1, as result an exponential smoothing time series model applied for predicting upcoming trend. However, the community level intervention R2a = 0.000 was significant (sig. 0.012) to use. ARIMA model for the prediction of upcoming trend.

Based on the appropriate time series modeling applied the anticipated trend on the level of interventions for child-focused social work services has forecasted for the period of 2024-2028. Fortunately all the four levels of intervention will probably demonstrate an increasing and stationary trend in a similar fashion like the previous periods (Significance test for all is > 0.05).

**Table 4.**
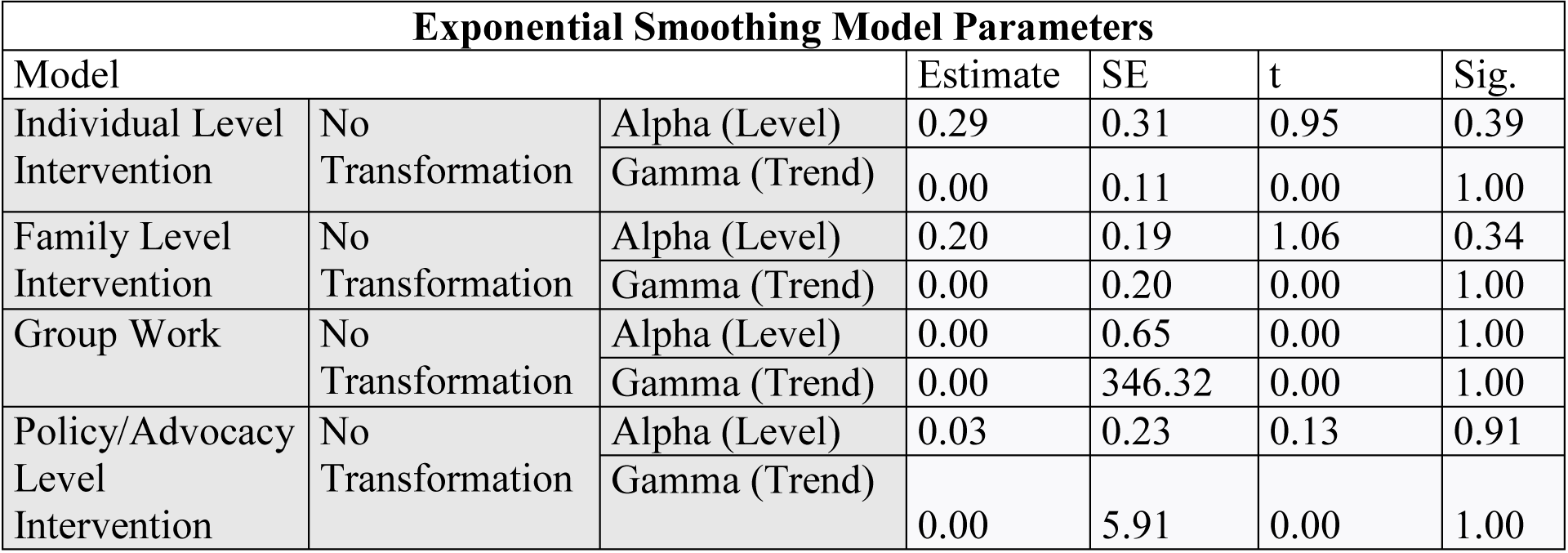
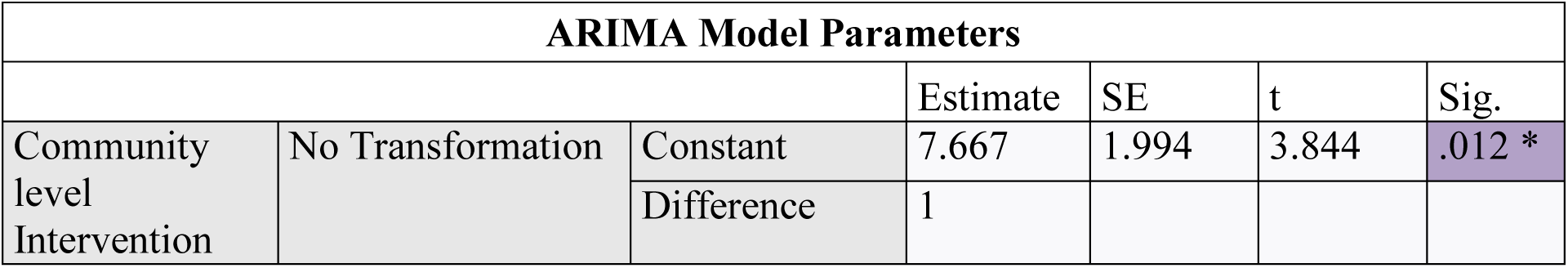
Parameters for time-series model levels of intervention.

| Exponential Smoothing Model Parameters |  |  |  |  |  |  |
| --- | --- | --- | --- | --- | --- | --- |
| Model |  |  | Estimate | SE | t | Sig. |
| Individual Level Intervention | No Transformation | Alpha (Level) | 0.29 | 0.31 | 0.95 | 0.39 |
|  |  | Gamma (Trend) | 0.00 | 0.11 | 0.00 | 1.00 |
| Family Level Intervention | No Transformation | Alpha (Level) | 0.20 | 0.19 | 1.06 | 0.34 |
|  |  | Gamma (Trend) | 0.00 | 0.20 | 0.00 | 1.00 |
| Group Work | No Transformation | Alpha (Level) | 0.00 | 0.65 | 0.00 | 1.00 |
|  |  | Gamma (Trend) | 0.00 | 346.32 | 0.00 | 1.00 |
| Policy/Advocacy Level Intervention | No Transformation | Alpha (Level) | 0.03 | 0.23 | 0.13 | 0.91 |
|  |  | Gamma (Trend) | 0.00 | 5.91 | 0.00 | 1.00 |

| ARIMA Model Parameters |  |  |  |  |  |  |
| --- | --- | --- | --- | --- | --- | --- |
|  |  |  | Estimate | SE | t | Sig. |
| Community level Intervention | No Transformation | Constant | 7.667 | 1.994 | 3.844 | .012 * |
|  |  | Difference | 1 |  |  |  |

**Figure 4.**
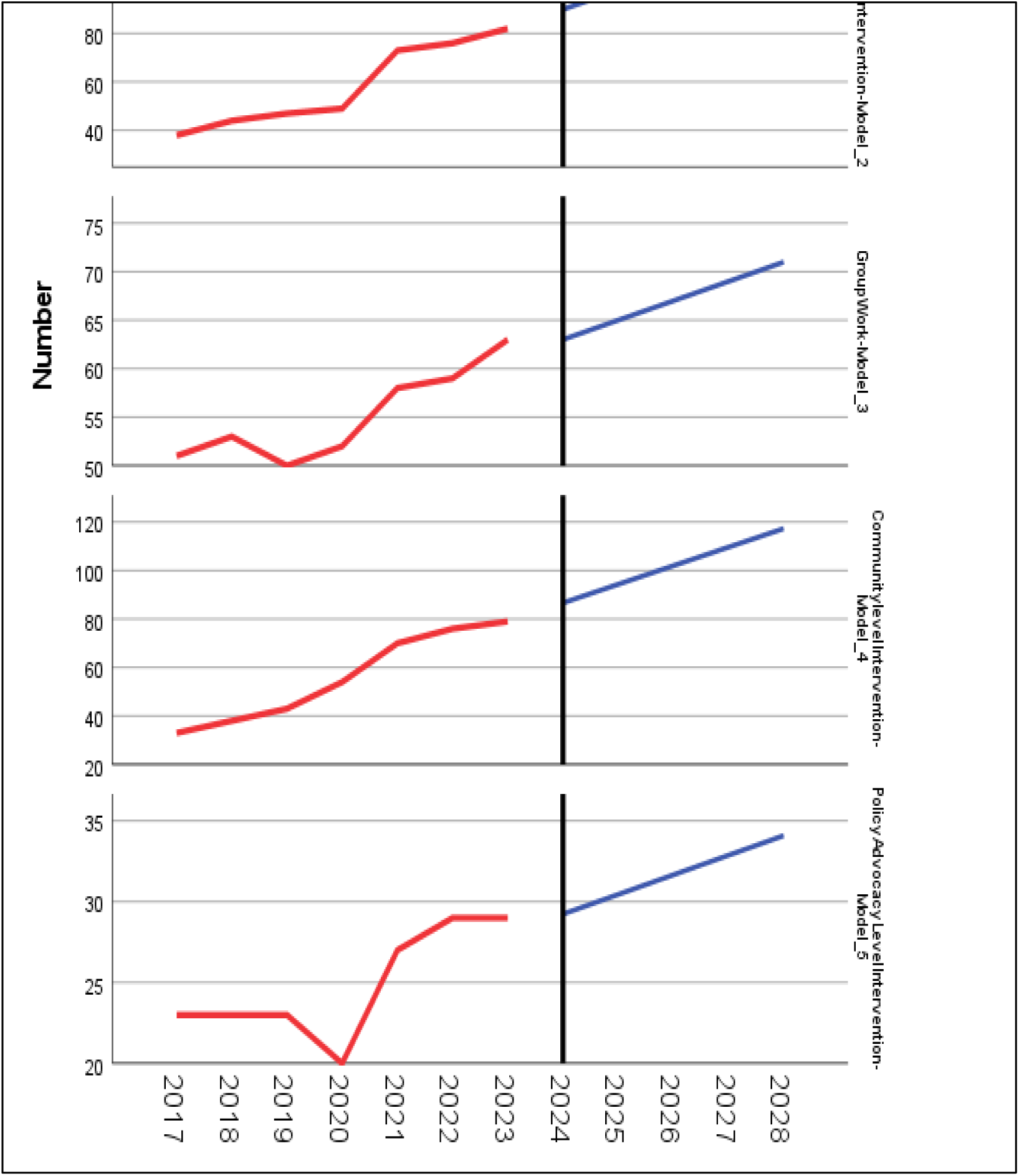
Forecasting on the levels of intervention for child-focused social work services.

To substantiate the result a chi-square test was employed to investigate the whether there is a significant association between the increasing trend of time and the frequency of child-focused social work services being provided by professional social workers. As per the cross tabulation in the below across the year 2017 – 2023 five top child focused social work services are mentioned by respondents that are being provided primarily by professional or trained social workers. These services involves (i) individual case management service, (ii) psychosocial support, (iii) community based child protection structure, (iv) alternative care arrangement, and (v) family tracing and reunification. However, the remaining 6 services are provided either by non-professional social workers.

Based on the result acquired from a quick sequencing chart it is found that (i) the average number of caseload per year showed non-regular (unpredictable) trend which shows in some years an increase and in other year a decrease. This is substantiated by the R-square result (R^2^ = - 0.09) which implies almost nil association with the increasing time period. (ii) the variable related to average number of children case one social worker manages per day shows a clear and stationary decreasing pattern from the year 2017 to 2023. The R-square (R^2^ = 0.878) result also infers a strong descending association of caseload per day with the increasing time period.

Moreover, a stationarized R-square test was applied to select an appropriate time series model to predict the upcoming 5 years trend (2024 up to 2028) in both average caseload per year and per day. As a result an ARIMA model was applied to run a time series analysis for the average caseload per year (R^2^_a_ = 0.313) and Exponential smoothing was applied to execute the time series analysis of average children caseload managed by one social worker per day (R^2^_a_ = 0.000).

The selected exponential smoothing and ARIMA models executed through considering the previous historical trend of both average refugee children caseload per year and day to forecast the trend for the years between 2024 and 2028. The forecasting result found out that (i) there will be an insignificant increasing trend in the number of average refugee children caseload until 2028, whereas (ii) the average number of refugee children to be managed (provided with child-focused service) will have a significant decreasing trend until 2028.

**Figure 5.**
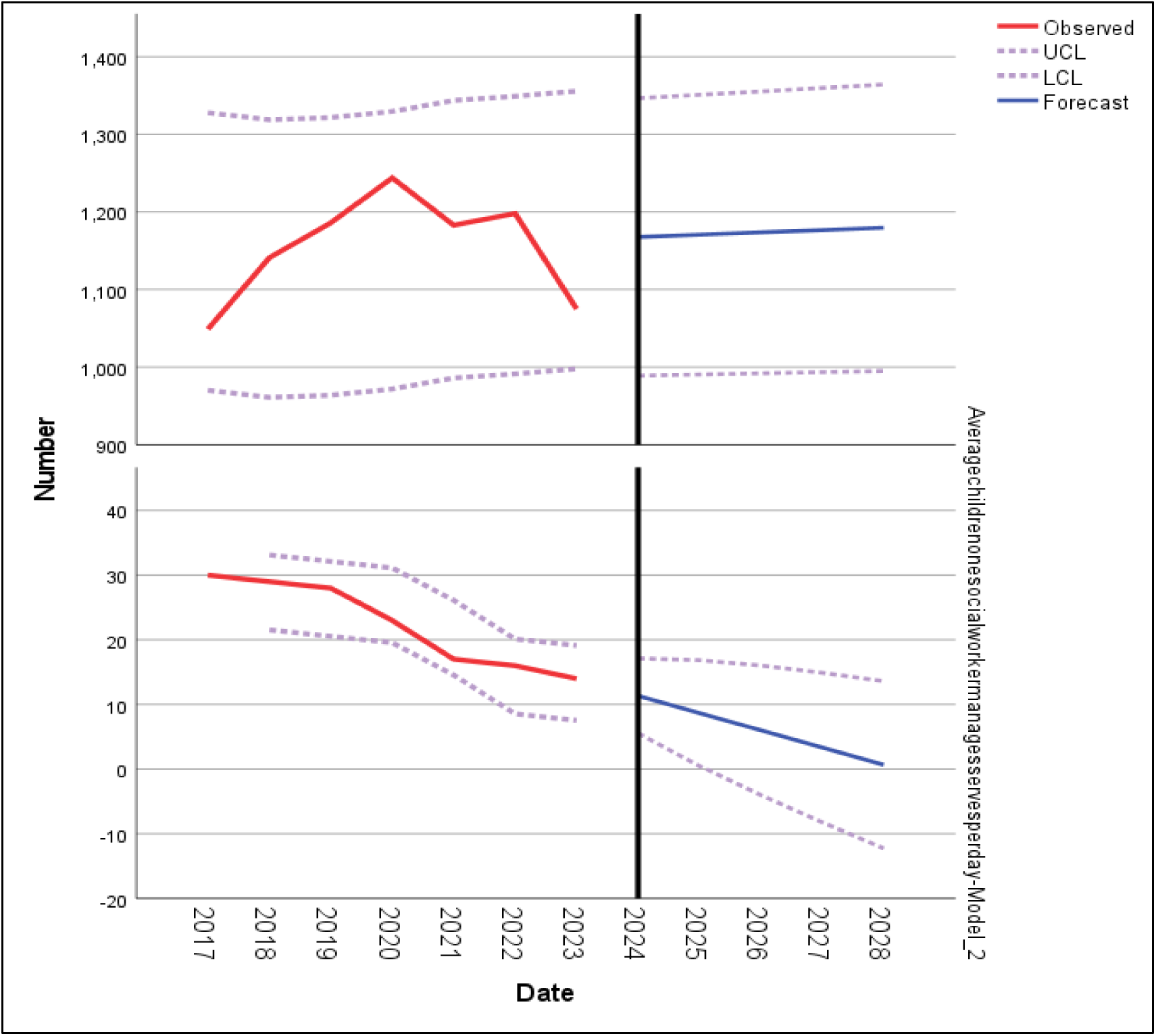
time series trend forecast of average caseload per year and day.

Complementary to the survey result few of focus group discussion (FGD) participants have mentioned that one social worker currently manages huge refugee children caseloads which is a fold of the previous year’s caseload.

## Discussions

The ultimate purpose of this chapter is concerned in providing a synthesis of the pressing findings acquired majorly from survey, at milder extent from key informant interview, and focus group discussion against to some of the theoretical and empirical perspectives highlighted in the literature review. For the purpose of easy articulation I have clustered the discussion in to four (4) categories aligning with the objectives of this study; (i) at the very primary level what natures do the studied organizations have concerning with the models of relationship between social work and organizations mentioned in the literature review, (ii) the trend in the type of child-focused social work services provided for refugee children as compared to standards in social work professionalism, (iii) how are the trends in the formats of delivering child-focused social work services progressing across the year 2017 through 2023, (iii) what are the pressing factors that plays a significant role in determining the trend in social work professionalism while providing child-focused social work services for refugee children

### Nature of the studied child-focused humanitarian organizations

Before heading to the discussion on the trend analysis component it is important to highlight the nature of the studied child-focused humanitarian organizations among the three Payne’s ‘Models of Relationship between Social Work and Organizations’ depicted in the literature review section. Findings infer that all the selected three child-focused humanitarian organizations follow a model suiting to ‘A managed relationship approach’. As a manifestation some of the findings complementing the above conclusion includes: (i) expectations from social workers to adhere to child-protection minimum standards in humanitarian action and other donor affiliated guidelines rather than social work professional standards (i.e.NASW), (ii) close to half of the social workers at refugee camp level are expected to spend their time in routine administrative and clerical activities instead of professional social work engagements, (iii) social workers are tasked to manage high caseload of refuge children which implies that the child-focused humanitarian organizations prioritize the cost effective management of huge caseloads with few number of social workers rather than considering how innovatively to manage the huge caseload without affecting the quality of child-focused social work services delivery, and (iv) due to the expectations of monitoring agencies for a fixed and uniforms monthly salary rate for social workers the studied child-focused organizations comply with this standard and forced to recruit unqualified social workers. Because of the collective efforts of the above findings, humanitarian organizations providing child-focused social work services for refugee children in Ethiopia do have a nature of managed relationship approach gives priority for the organizational objectives, regulations and some of the commitments made to authorities, donors and public.

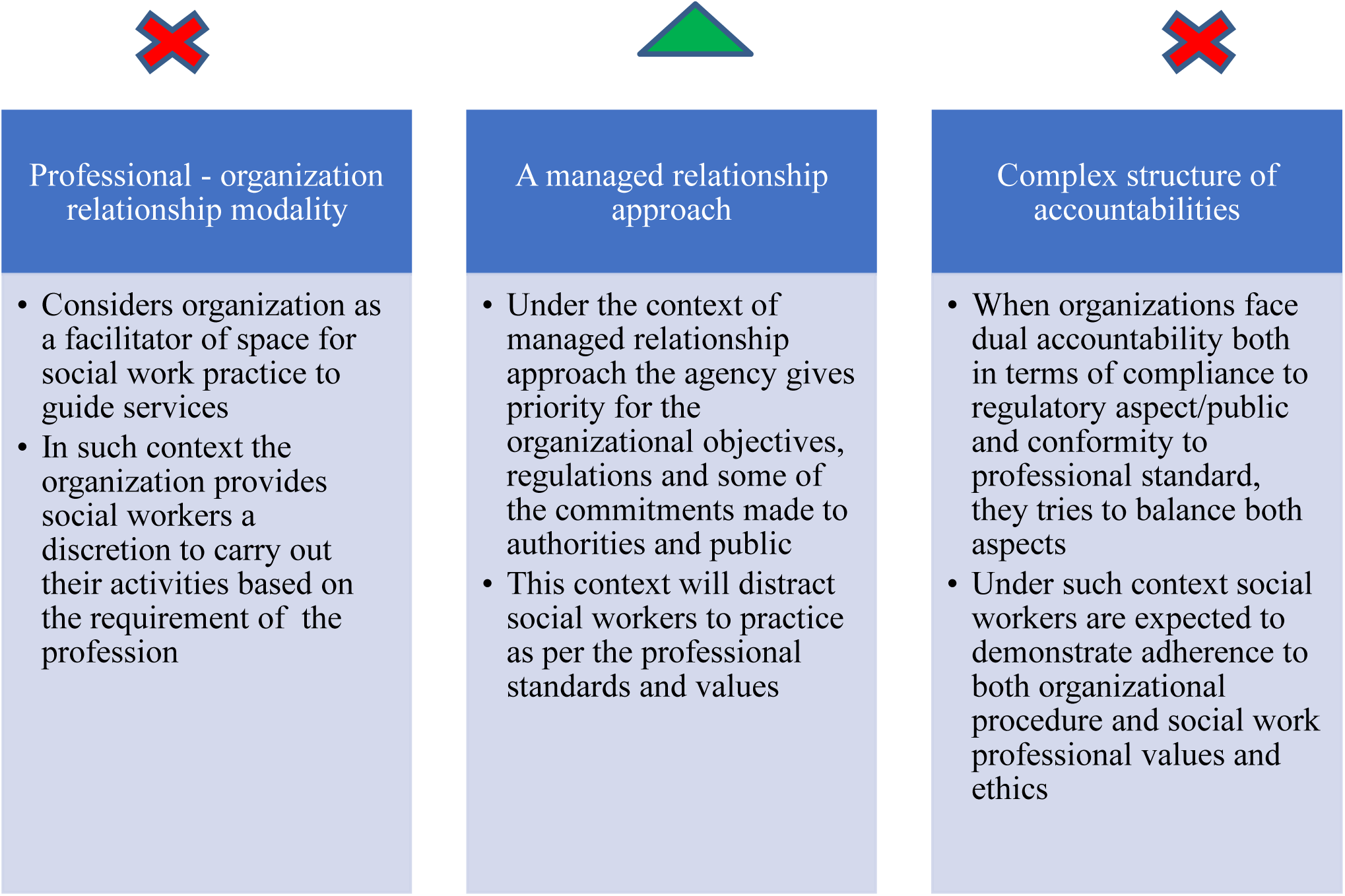

### Trend in type of child-focused social work services

This study has managed to map out 11 (eleven) types of child-focused social work services which has been provided to refugee children by humanitarian organizations in Ethiopia since 2017. However, the most interesting finding is the eleven service types can be categorized in to three major categories of service and trend natures across time.

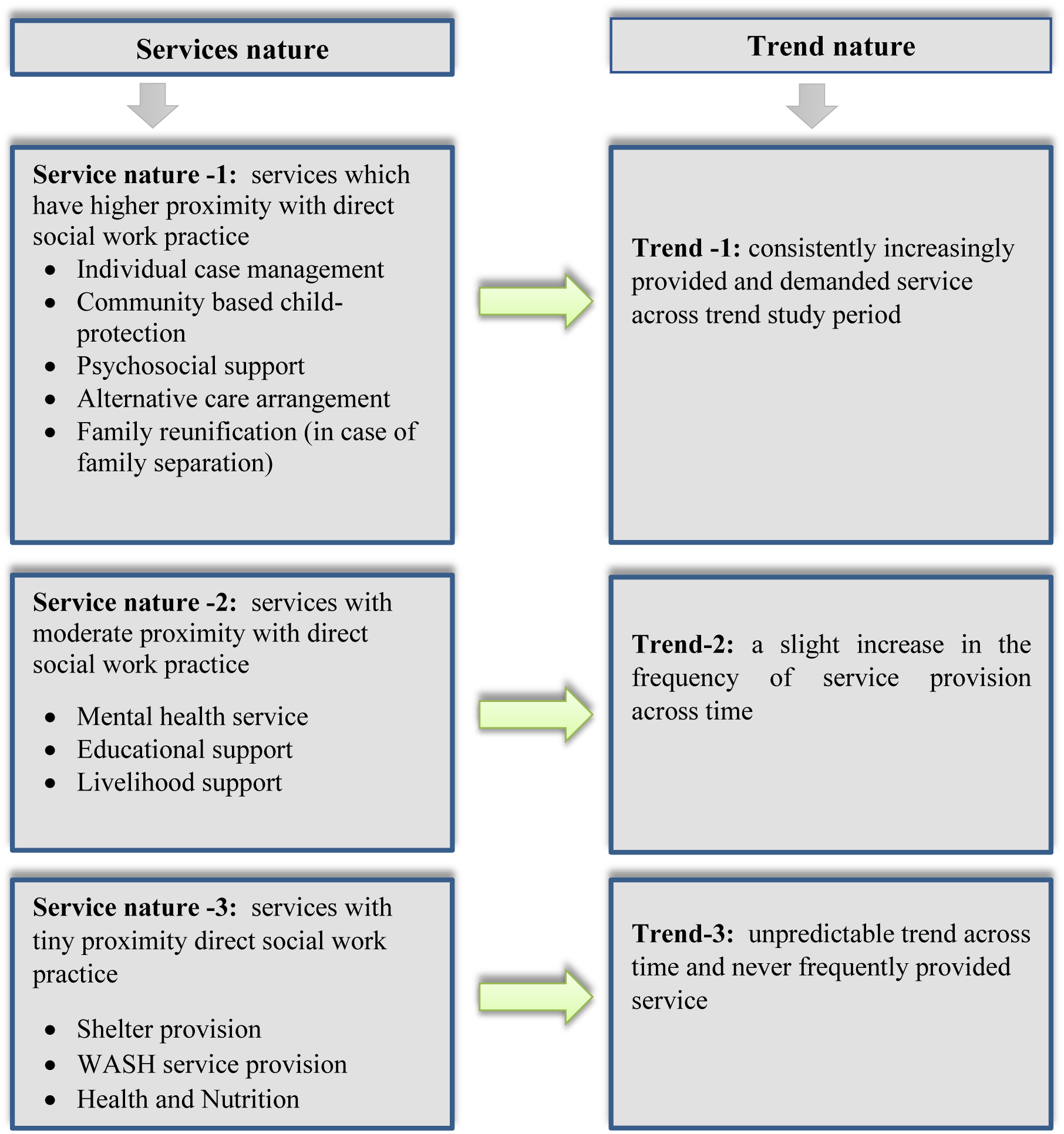

Additionally, the Child Protection Minimum Standards (CPMS) in humanitarian action framework is becoming the leading standard for service delivery, demonstrating a strong alignment with the NASW standards which is the fundamental frame work in Social work practice. Ethical conduct is also receiving greater emphasis, with organizations implementing safeguarding policies and codes of conduct. Some social workers even go beyond by utilizing the NASW standards for social work practice.

Based on the desk review conducted as part of this study the researcher has identified several alignments between CPMS in humanitarian action and NASW standards, as manifestation three crucial areas of alignments has been identified.

#### Core Values

Both the CPMS and NASW standards emphasize core values central to social work, such as the dignity and worth of the person, the importance of human relationships, and the pursuit of social justice.

#### Client Centred Approach

Both frameworks advocate for a client-centered approach, ensuring services are tailored to meet the specific needs and vulnerabilities of each child.

#### Empowerment (Egalitarian relationship between client and social worker)

Both CPMS and NASW standards promote the empowerment of children and families, encouraging their participation in decision-making processes.

#### Ethical Conduct

Both frameworks provide guidance on ethical conduct; ensuring social workers uphold professional integrity and act in the best interests of their clients.

Hence, the increasing use of both CPMS and NASW standards, particularly among social workers with academic training in social work or related fields, signifies a positive trend. This demonstrates a growing commitment to integrating best practices in child protection with a strong ethical foundation for social work practice. However, these positive trends are countered by significant challenges that hinder the effectiveness of service provision. The most concerning factor is the overwhelming workload faced by social workers. The dramatic increase in caseloads, with a single worker now managing 100-150 children compared to the previous 50-57, severely limits the time and attention they can devote to each case. This inevitably impacts the quality of services offered and the ability to adhere to professional standards. Moreover, it bypasses the CPMS standard concerned with a 1:25 social worker per day as per ***Standard 18: 18.1.7.*** (Alliance for Child Protection in Humanitarian Action, 2019). Compounding this problem is the lack of readily available updated training for social workers. Without on-going professional development, it becomes difficult for them to stay abreast of the latest best practices and refine their skills. While a slight decrease was observed in staffing shortages, concerns remains regarding the qualifications of the personnel hired due to monthly incentive payment ceiling set by government counterpart on refugees(RRS) and UNHCR which hindered the motivation of social workers and affected also the quality of social work services as per the minimum standards. Security concerns are also on the rise, creating an additional layer of stress and potentially hindering the delivery efforts of child-focused social work services. Based on the observation made by the researcher also the on-going tension between the refugee and surrounding host community has been restricting movements of child-protection staffs to refugee camps from the organizations coordination offices based at the capital of respective regions (i.e. Gambella and Afar regions) which forces the incompetent refugee incentive social workers to unsupervised for several days.

### Trends of child-focused service delivery formats

One of the most significant trends identified is the growing emphasis on needs assessments as a basis for the selection of child-focused social work services. This increasing use of needs assessments indicates the growing commitment of those child-focused humanitarian organizations to apply a client-centred approach and ultimately ensure most pressing protection concerns of refugee children. However, it is found that the need assessments consult majorly the caregivers (family of the refugee children) and other adult members. This totally ignores the consultation with refugee children themselves which contradicts with one of the child protection principles in humanitarian action that is *‘child participation’*.

Another positive trend is the increasing collaboration among child-focused humanitarian organizations. The collaboration for service delivery and knowledge sharing is significantly rising across time this will contributes to in place a more holistic approach and potentially helps also synergy between child-focused humanitarian organizations which will have dual implication on the quality delivery of child-focused social work services and helps to tackle the resource gap observed which led the social workers to handle huge caseload. Additionally, of daily supervision social workers engagement by project officers serves as a quality control measure, ensuring adherence to established standards.

However, a trend which requires special attention has also emerged. For instance, while social workers in the sampled organizations collects feedback from various segments of the refugee population towards how child-focused services have been provided to refugee children, a very little attention was given to collect feedback from the children themselves. This could indicate communication gaps between social workers and children, or a hesitation on the part of children to express their concerns and violets their right embedded in the United Nations Child Right Convention (UNCRC) which is their right to participation. Further exploration is needed to understand the reasons behind this decrease and ensure children’s voices are adequately heard and incorporated into the child-focused social work service delivery process for refugee children.

### Conclusion and Recommendations

As per the findings common services include child protection, psychosocial support, family reunification, and education. There’s a rise in individual case management, alternative care, and community-based protection services. Child-friendly spaces are the preferred location for delivering these services. Social workers are increasingly using individual, family, and community-level interventions. Challenges include high caseloads, resource limitations, and ethical dilemmas in prioritizing services. Positive trends include using the Child Protection Minimum Standards framework, a more client-centered approach, and increased collaboration with families. Moreover, the education status of the existing social workers infers that majority of them are high school complete which will potentially affect the quality of the child-focused social work service being provided for refugee children.

Considering the findings of the study participation of children is crucial for ensuring child-focuses social work services and is relevant to meet their needs. Develop effective mechanisms to lobby feedback from the refugee children directly such as child-friendly surveys and focus group discussion with the children themselves needs to be given emphasis. There should also be a feasible and innovative modality of high caseload management strategies to tackle the challenge of decrease in the number of caseworks due to funding limitations.

## Data Availability

All the raw data used to produce the study are available at hand and can be found from Addis Ababa University Digital Library

## Notes

### Competing Interest Statement

The authors have declared no competing interest.

### Funding Statement

This study is self sponsored as part of my thesis for acquiring my Masters of Arts in Social Work

### Author Declarations

Addis Ababa University School of Social Work

### Summary of Updates

I am revising for the second time the name of Corresponding Author in the Author list section from 'Mesele Mengstaeb' to 'Mesele Mengsteab' with spelling error in father name.

## References

ACSO. (2021). List of Foreign registered CSOs in Ethiopia. Addis Ababa: FDRE Civil Societies Organization.

Alliance for Child Protection in Humanitarian Action. (2019). Child Protection Minimum Standards in Humanitarian Action. Alliance for Child Protection in Humanitarian Action.

Alliance for Child Protection in Humanitarian Action. (2023). The Evolution of the Child Protection in Humanitarian Action Sector. Alliance for Child Protection in Humanitarian Action.

Antoni, A., Reinecke, J., & Fotaki, M. (2023). Making Time to Care, and Caring for Time: ‘Tricking Time’ to Cope with Conflicting Temporalities in a Child Protection Agency. Journal of Business Ethics, 645–663.

Austin, D. M. (2002). *Human Service Managment - Organizational leadership in Social Work Practice*. New York: Columbia University Press.

Balgopal, P. R. (2000). *Social Work Practice with Immigrants and Refugees*. New York : columbia university press.

Barnett, M. (2013). *Emipre of Humanity : A History of Humanity*. New York: Cornell University Press.

Bill Tassie, e. (1998). Evaluating Social Service Agencies: Fuzzy. Voluntas: International Journal of Voluntary and Nonprofit Organizations*, Vol.* 9, No. 1, 1, 59–78.

Blau, P. M. (1965). The Comparative Study of Organizations. SAGE ILR Review, 323–338.

Chlebanova, L., & Rusnakova, M. (2022). Organizational culture in social services focused on the workers’ well-being. SHS Web of Conferences 131, 03003. 131. p. 10. Slovakia: EDP Sciences.

Claiborne, N. (2004). Presence of Social Workers in Nongovernment Organizations. *National Association of Social Workers*, Inc, 215.

Datta, S. S. (2017). Role played by social workers in Non Governmental Organizations in preventing human trafficking in India. International Journal of Humanities and Social Science Invention, 34–37.

Edmos, M., & Munyaradzi, M. (2020). In search of culturally sensitive social work practice in the care and protection of children in Zimbabwe: the case of Remba/Lemba culture. AJOL African Journals, 10(2), 93–100.

Engel, G. V. (1970). Professional Autonomy and Bureaucratic Organization. Administrative Science Quarterly, 15(1), 12–21.

Engstrom, S. (2014). Organizational justice and child protection Social Work within the Child Protection function of Scottish Local authorities. J. Workplace Rights, 17(3-4), 347–366.

European Union, E. (2012). Time Series Analysis. European Union.

Fuller, W. A. (2009). *Sampling Statistics (Wiley Series in Survey Methodology)*. Iowa State University: A John Wiley & Sons, Inc..

G.Tornatzky, L. (1979). Dimensions of Interorganizational Interaction In social Service agencies. Journal of Community Psychology, 198–209.

Greene, G. (1990). Autonomy and Professional Activities of Social Workers in Hospital and Primary Health Care Settings. Health and Social Work.

Hailu, D. (2013). The development and management of social workers in Ethiopia: A situational analysis. International Journal of Social Welfare, 392–401.

Harvard Humanitarian Initiative. (2024). Retrieved from Children in Crisis: https://hhi.harvard.edu/children-crisis

Hasenfeld, Y. (2010). *Human Services as Complex Organizations*. Los Angeles: SAGE Publications, Inc.

Hillel, R. S. (2023). Decolonizing Mental Health and Psychosocial Support (MHPSS) Interventions in the Humanitarian System. Journal of Mental Health and Psychosocial Support in Conflict-Affected Areas, 20–29.

Hiwot, T. (2016). Social Work Practice: Roles and Challenges of Social Workers in Selected Public Hospitals in Addis Ababa. Addis Ababa University Institutional Repository.

Hjärpe, T. (2020). Measuring Social Work Quantity as Quality in the Social Services. Statsvetenskaplig tidskrift, 23–45.

I-TECH. (2010). *Assessment of social work preservice training institutions in Ethiopia - A report from the Strengthening the*. Addis Ababa : I-TECH.

Junghee Bae, e. (2019). Compassion Satisfaction Among Social Work Practitioners: The Role of Work–Life Balance. Journal of Social Service Research.

Kebede, W. S. (2019). *Practice and Challenges of Child Protection by NGOs: The Case of Save the Children at Jewi Refugee Camp, Gambella, Ethiopia*. Addis Ababa: St. Mary’s University.

Leighninger, L. (1978). Professionalism and Social Work Education: Substance and. The Journal of Sociology & Social Welfare*, Vol.* 5(2), 188–213.

Lenka Chlebanova, a. M. (2022, January 11). Organizational culture in social services focused on on the workers’ well-being. SHS Web of Conferences, 133.

Meagherb, G.. & Healya, K. (2007). Social Workers’ Preparation for Child Protection: Revisiting the Question of Specialization. Australian Association of Social Workers, 321–335.

Melaku, G. (2022). Decolonizing Social Work Practice: A Case from Ethiopian Refugee Settings. Journal of Social Service Research, 273–284.

Morris, R. (1974). The place of social work in the human services. Oxford Journals, 519–531.

Mugumbate, J., & Nyanguru, A. (2015). Exploring African philosophy: the value of ubuntu in Social Work. AJOL African Journals Online, 3(1(2013)), 82–100.

NASW. (2011). Social Workers in Social Services Agencies. Occupational profile.

NASW. (2021). code of ethics. Retrieved December 18, 2023, from National Social Worker Association: https://www.socialworkers.org/About/Ethics/Code-of-Ethics/Code-of-Ethics-English

Nordesjö, K. (2020). Framing Standardization: Implementing a Quality Management System in Relation to Social Work Professionalism in the Social Services. *Human Service Organizations: Management*, Leadership & Governance, 44(3), 229–243.

O’Connor, M. K., &, F. N. (2009). *Organization Practice: A Guide to Understanding Human Service Organizations* (2nd ed.). New Jersey: John Wiley & Sons, Inc.

Okitikpi, T., & Aymer, C. (2003). Social work with African refugee children and their families. Child and Family Social Work, 8, 213–22.

P. Ž., J. V., & R. A. 2018. Philosophy and Paradigm of Scientific Research. Intech Open, 121–139.

Payne, M. (2006). *What is professional social work?* (J. Campling, Ed.) University of Bristol, Great Britain: Policy Press.

Peter, M.-S. (1985). *International Humanitarian Assitance : Disaster Relief Actions in International Law and Organization*. Netherlands: Springer.

Pumima George, e. (2007). “PROVIDING SERVICES”: Voices of Service Users on Structural Social Work Practice in Community-based Social Service Agencies. Canadian Social Work Revie, 5 - 22.

Rahel, A. (2017). Social Work Practice in Institutional Childcare System: The Case of Governmental Childcare. Addis University Repository.

Rich Furman, M. G. (2021). Navigating Human Service Organizations, Ren Winnett. New York : Oxford University Press.

Schopper, C. E. (2022). Organizations Involved in Humanitarian Action: Introducing a New Dataset. International Studies Quarterly.

Shin, Y. (2017). *Time Series Analysis in the Social Sciences*. Oakland: University of California Press.

Singh, K. (2007). *Quantitative Social Research Methods*. New Delhi : Sage Publication.

T.F. Leung, T. (2010). Social work professionalism in self-help organizations. International Social Work, 53(4), 474–488.

Triggs, S. (2020). Making a Difference Again: How Using Coaching Enabled Children’s Social Workers to Enhance their Practice & Fulfil Their Vocational Aspirations. International Journal of Evidence-Based Coaching and Mentoring, 77-87.

UNHCR. (2020). *Camp Profile Nguenyyiel Refugee Camp*. UNHCR.

UNHCR. (1951). Convention Relating to the Status of Refugees.

UNHCR. (1993). *Policy on Refugee children*. UNHCR.

UNHCR. (2014). *UNHCR Operational Report*. Addis Ababa: UNHCR.

UNHCR. (2020). *Ethiopia Operation 3W Dashboard*. UNHCR.

UNHCR. (2021-2025). *Refugee and Protection Strategy*. Ethiopia: UNHCR.

UNHCR. (2023). Overview of Refugees Camps and Sites. Ethiopia: UNHCR.

UNHCR. (2023). *Refugees and Asylum-seekers Statistics*. Ethiopia : UNHCR.

Vibha, K., & A. Walsh, C. (2019). Pragmatism as a Research Paradigm and Its Implications for Social Work Research. Social Sciences Journal.

Wicleek, N. K. (2020). From a Refugee Camp in Ethiopia as a Social Worker to Working with Refugees as an Expert Psychotherapist in Ethiopia: A Story of a South Sudanese Canadian Immigrant. Journal of Mental Health and Psychosocial Support in Conflict Affected Areas, 18(2), 172–175.

Wuenschel, P. C. (2008). The Diminishing Role of Social Work Administrators in Social Service Agencies. Central Michigan University, 5–17.

